# Novel non-invasive physical photobiomodulation can treat congenital colour vision deficiency and enhance color vision recognition ability: a randomized, single blind, controlled clinical trial

**DOI:** 10.1101/2024.08.13.24311912

**Authors:** Nandi Bao, Liang Jia

## Abstract

**Background:** Color vision deficiency (CVD) is a common congenital ophthalmic disease, and there are no effective therapeutic measures currently available for this patient population. This study aimed to explore the efficacy of photobiomodulation (PBM) in CVD patients.

**Methods:** A randomized controlled trial design was applied in this study, whereby 80 patients with red-green CVD were randomly divided into PBM treatment and control groups. Color vision was assessed after 1, 2 and 4 weeks of treatment. Statistical analysis of treatment was carried out using the Kruskal-Wallis test with Dunn’s test for multiple comparisons using SPSS and R software.

**Findings:** Compared to the control group, the color vision of CVD patients exhibited a significant improvement in the PBM treatment group for different parameters (P_s_□0.05).

**Interpretation:** PBM therapy could significantly improve color vision in patients with congenital CVD, especially in patients with green CVD.

## Introduction

Color vision deficiency (CVD) is a sex-related genetic disease that reduces the perception of a wide range of colors, of which red-green CVD is the most frequent in humans^[1, 2]^. Besides genetic therapy, which has not yet been applied in clinical practice, there is currently no effective treatment of CVD in humans^[3, 4]^. Photobiomodulation (PBM) is a low-intensity/laser-assisted clinical therapy based on non-ionizing electromagnetic radiation (mainly composed of visible light and infrared), which exhibits anti-inflammatory properties and promotes wound healing. PBM is widely used in the treatment of many diseases, such as acute myocardial infarction, Alzheimer’s disease, Parkinson and other neurodegenerative diseases^[5]^. The mitochondrial enzyme cytochrome C oxidase is a key photoacceptor of light^[6]^, and the beneficial effects of PBM include enhanced mitochondrial energy generation through increased RNA and protein synthesis^[7]^. Markowitz et al. showed that PBM could be used to treat refractory macular degeneration^[6]^. An intervention study of 24 healthy people aged 28 to 72 found that blue visual function was significantly improved in the over-40 age group after exposure to 670 nm light for 3 minutes per day for 2 weeks^[8]^. These positive clinical findings, coupled with PBM’s known mitochondrial-based mode of action, suggest that PBM of different wavelengths may improve the colour visual function of patients with CVD. To validate this hypothesis, we conducted a 4-week PBM intervention study for red/ green/ red-green CVD patients of different ages, which provided new and effective clinical evidence for the treatment of congenital colour vision abnormalities.

## Subjects and methods

### 1 Study subjects

In this single-center study, a randomized controlled design was carried out. 80 patients at the Chinese PLA General Hospital, Beijing, China, who met the inclusion criteria and did not meet the exclusion criteria, were recruited by posting posters and advertising online on social media.

#### 1.1 Inclusion and exclusion criteria

Inclusion criteria: (1) Diagnosis of congenital CVD (including colour blindness and colour weakness) established by professional ophthalmologists; (2) Patients with normal or corrected-to-normal visual acuity of 20/25 or better; (3) Patients that agreed to participate in the study and signed the informed consent. Patients were excluded for the following reasons: (1) Diagnosed with other eye diseases that affect vision and colour vision, such as glaucoma, cataracts, optic neuritis and acquired colour vision abnormalities; (2) Diagnosed with mental illness and mental abnormality; (3) Diagnosed with other serious comorbidities (acute myocardial infarction, advanced cancer, etc.).

#### 1.2 Ethics and trial registration

This study was approved by the Ethics Committee of PLA General Hospital (Ethical Batch Number: KY2021-017) and registered on the Chinese Clinical Trial Registry website (Registration Number: ChiCTR2200056761). All included subjects provided informed consent. All experimental protocols were performed in accordance with the guidelines provided by the committee approving the experiments and with the Declaration of Helsinki.

### 2 Experimental methods

#### 2.1 Method of randomization and grouping

Simple randomization was adopted in this study. Randomization was performed with computer-generated random numbers, and participants were randomly divided into 2 groups in a 3:1 ratio: treatment group (n=60) and control group (n=20). The diagnosis of CVD was mainly based on medical history, clinical examination and the Farnsworth Munsell 100 (FM-100) Hue colour vision test. According to the diagnosis, the subjects in the treatment group were divided into treatment group 1 (red vision abnormality patients, n=17), treatment group 2 (green vision abnormality patients, n=21) and treatment group 3 (red-green vision abnormality patients, n=22).

#### 2.2 Colour vision function test methods

##### 2.2.1 Fm-100 Hue test

The Fm-100 Hue test detection program (https://www.colour-blindness.com/farnsworth-munsell-100-hue-colour-vision-test/) was used to obtain the total error score (TES), the type of CVD and the severity of CVD. TES is a widely acknowledged parameter for the quantitative evaluation of CVD; the higher the TES, the more serious the CVD.

##### 2.2.2 Ishihara’s colour blindness test

Patients were asked to identify all numbers on the color blindness test cards in Ishihara’s colour blindness book. Each correct answer scored 1 point, leading to a total score ranging from 0 to 20.

##### 2.2.3 Colour Blind Check (CBC) test

The smartphone app “Colour Blind Check” (https://play.google.com/store/apps/details?id=ch.colblindor.Colourblindcheck) was used to test the patients, and the total score of CBC, the severity of CBC and the type of CBC (red/ green/ blue colour vision deficiency) were obtained.

##### 2.2.4 Yu Ziping’s colour blindness test

Patients were asked to select the 2nd-16th, 3rd-40th and 42nd cards in Yu Ziping’s colour blind book (6th edition) and identify all numbers on the colour blindness test card. Each correct answer yielded 1 point, leading to a total score ranging from 0 to 20.

#### 2.3 Treatment for CVD

In treatment groups, patients in groups 1, 2 and 3 were treated with different wavelengths of light while wearing VR glasses (a smartphone assembled on a VR box compatible with smartphones)(Storm Mirror Litte D, Beijing Fengfeng Magic Mirror Technology Co., Ltd). Pictures of the PBM treatment and VR glasses used are provided in the appendix (supplementary appendix 1). Patients in treatment group 1 received red light irradiation (λ = 621 nm, E = 2.02 lx, E = 8.69 * 10-3 w/M2, CCT = 1001 K) ; patients in treatment group 2 received green light irradiation (λ = 524nm, E = 12.68 lx, EE = 2.21 * 10-2W/m2, CCT = 6257K) ; in treatment group 3, patients were treated with red light and green light alternately (the parameters of irradiation were the same as those in treatment groups 1 and 2). The duration of irradiation per session was 6 minutes and 40 seconds twice a day (morning and evening, with an inter-block period of 12□hours), lasting for 4 weeks. All participants were required to be awake and blink normally during irradiation therapy.

Patients in the control group did not receive any treatment. All participants were asked to undergo the FM-100 Hue test, Ishihara’s colour blindness test, CBC test and Yu Ziping’s colour blindness test before and at the end of week 1, week 2 and week 4 of treatment (Figure 1). A CONSORT diagram illustrating the study protocol is provided in Figure 2. Schematic diagram of color vision tests are shown in the supplementary appendix 2.

**Figure1:**
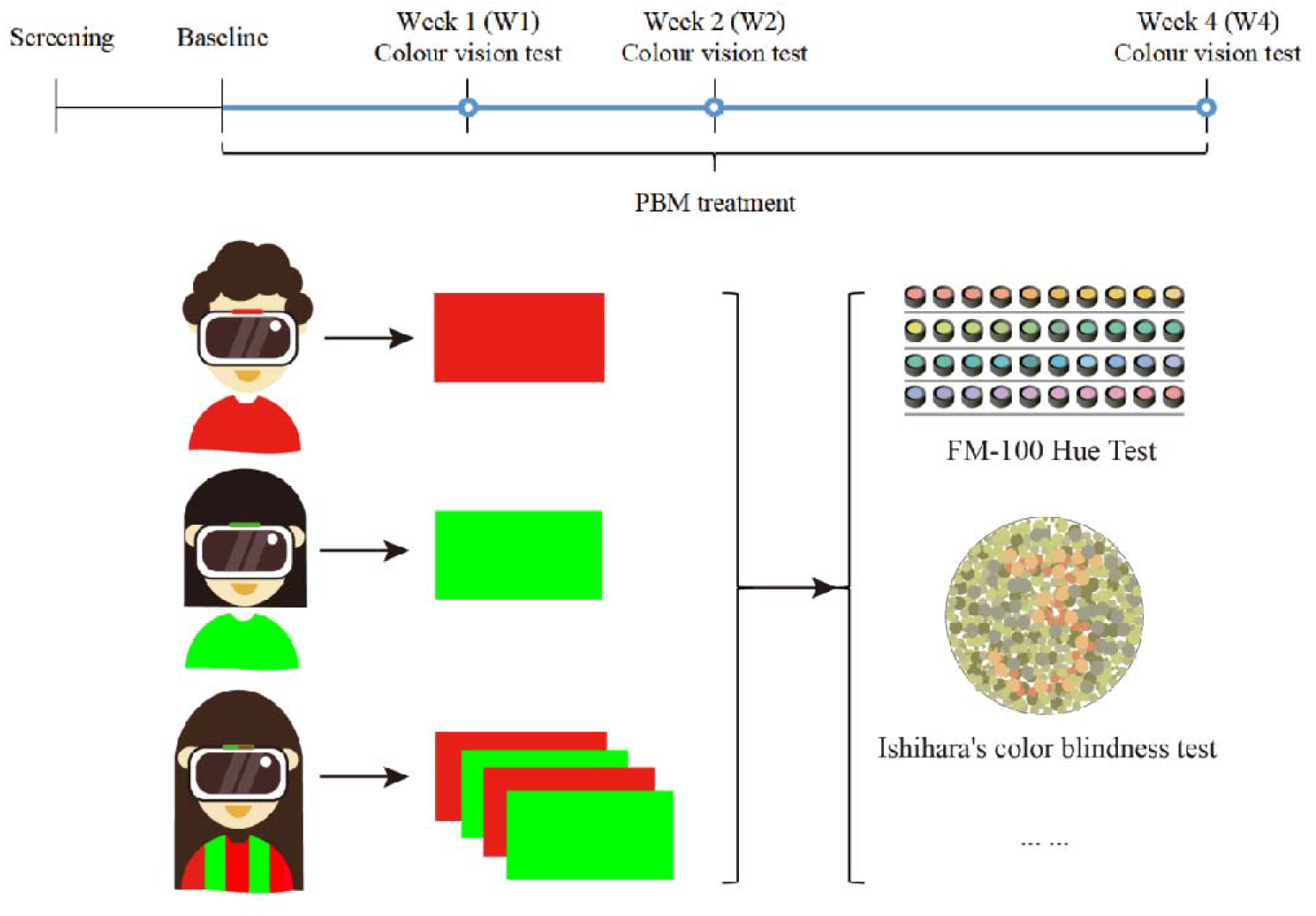
Diagram illustrating the clinical study design. Subjects who met the inclusion/exclusion criteria were included in the study and underwent 4 weeks of PBM treatment. The colour perception was assessed at the end of the 1st, 2nd and 4th week of treatment. Subjects in the treatment group were divided according to the types of CVD and received corresponding PBM treatment.

**Figure2:**
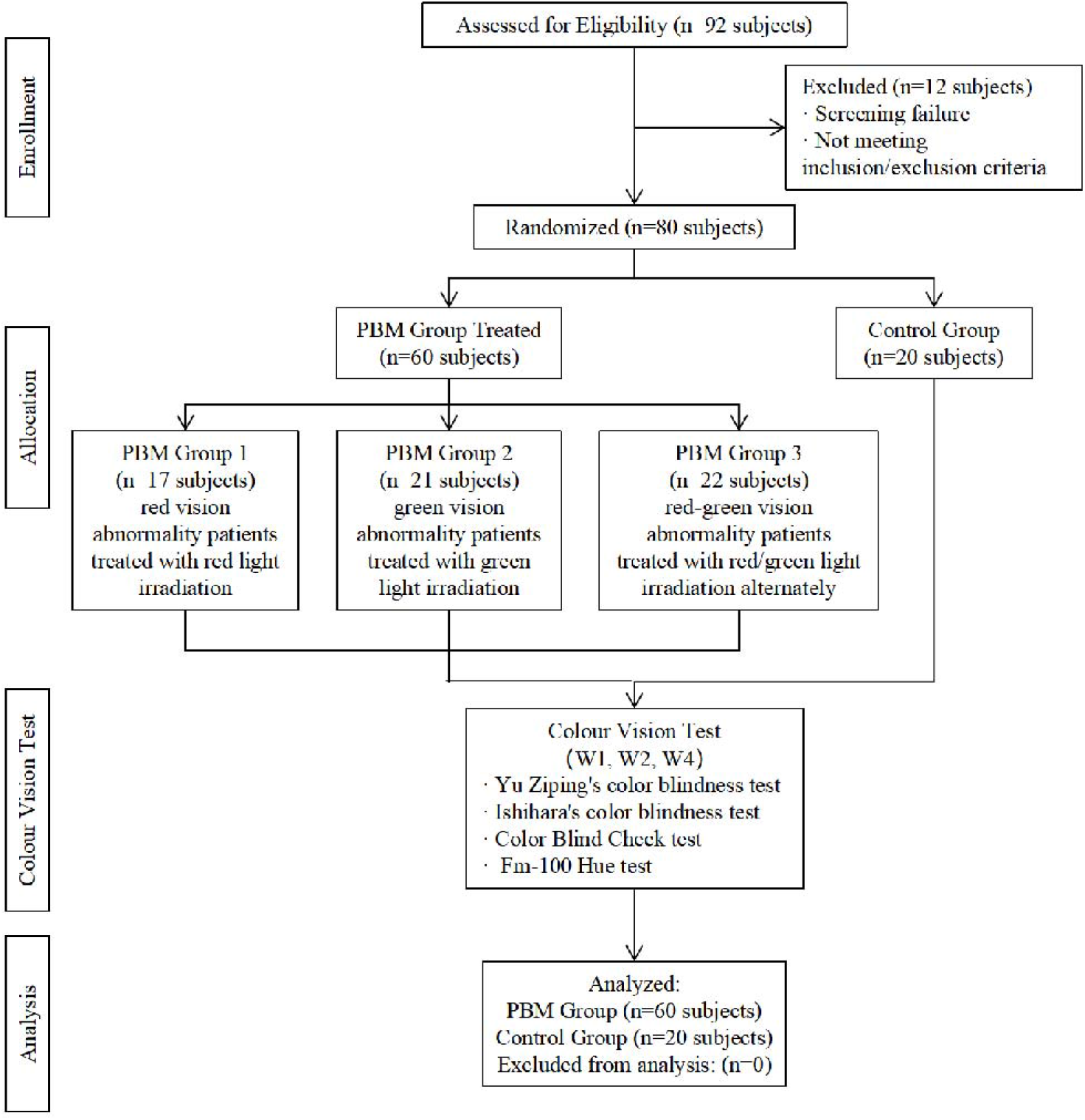
Consort diagram of treatment for different groups during each study phase.

**Figure3:**
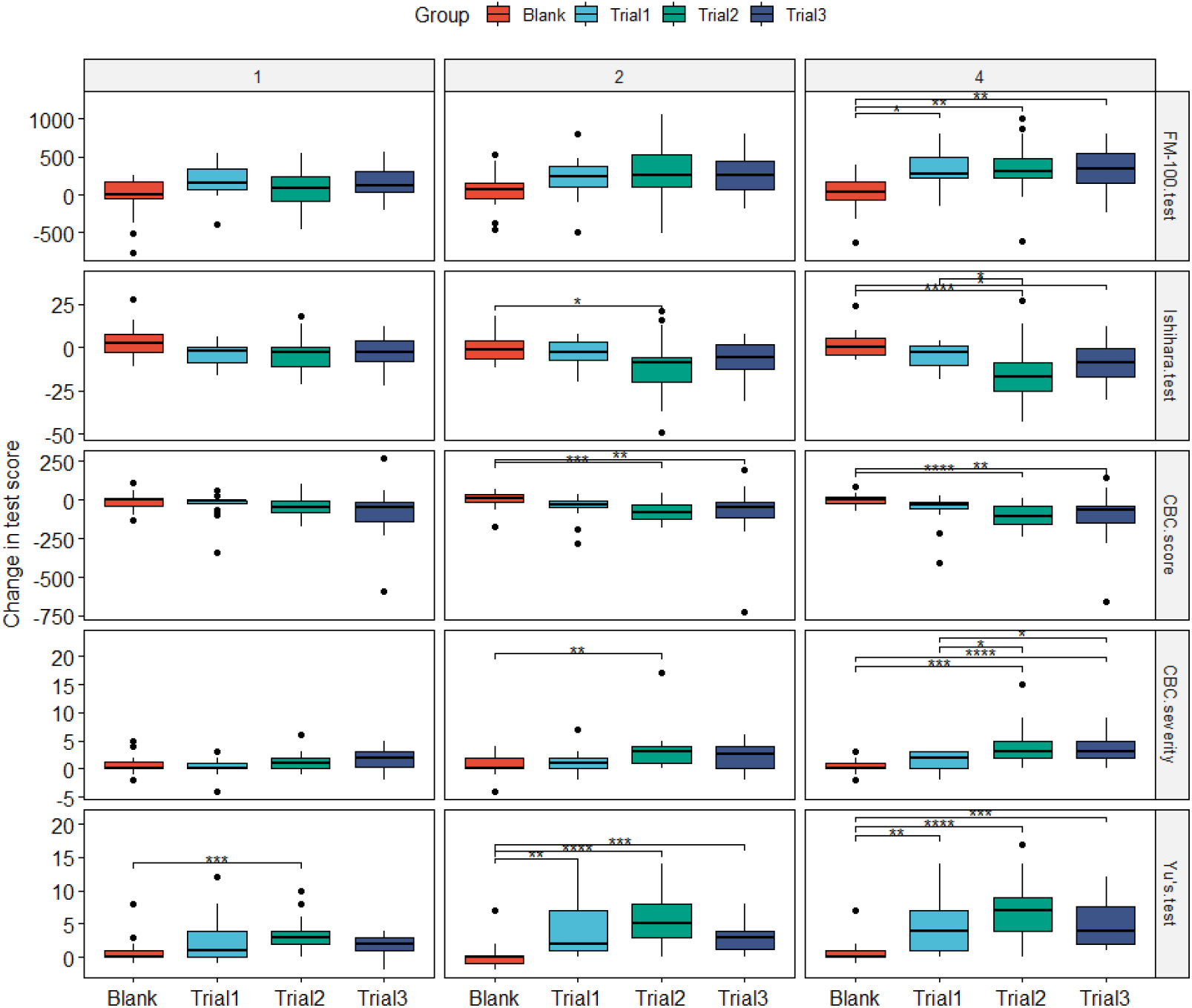
Change in the results of different evaluation methods after 1, 2 and 4 weeks between the treatment and control groups. * p□0.05, **p < 0.01, ***p < 0.001 and ****p < 0.0001. Kruskal-Wallis test, followed by Dunn’s multiple comparison test for between group comparisons.

**Figure4:**
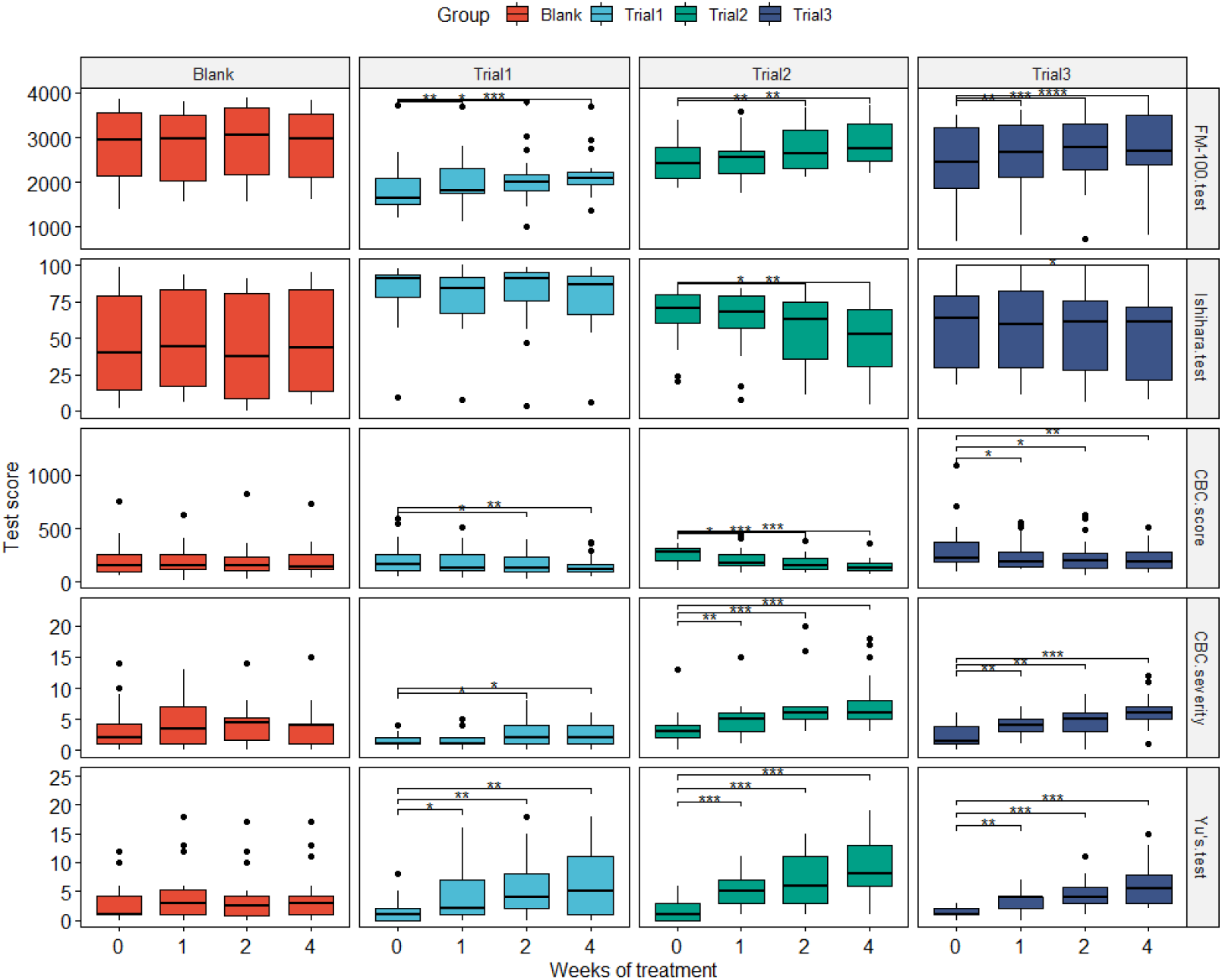
The results of different evaluation methods in treatment groups 1, 2, and 3 and the control group at baseline and after 1, 2 and 4 weeks of treatment. * p□0.05, **p < 0.01, ***p < 0.001 and ****p < 0.0001. Friedman test followed by Wilcoxon matched-pairs signed-rank test for pairwise comparisons. The medians and IQRs of different evaluation methods in each group are available in supplementary file. 4 Safety/adverse events. There were no adverse events over the study period.

#### 2.4 Sample size estimation

In this study, the test level is set to α=0.05□Z1-α/2=1.96□ and the test efficacy is 1-β=0.90□Zβ=1.28□.According to the following formula:

d is the difference of the mean between the two groups, σ is the standard deviation. The main outcome of this study is Fm-100 Hue test score. According to the pre-experimental results, d=0.14 and σ=0.12, and after calculation, the sample size of each group is 15. Considering the possible loss of follow-up of the subjects (with a lost to follow-up rate of about 10%), it is estimated that the sample size of the experimental group is 60. According to the 1:3 matching principle, the sample size of the control group was 20.

#### 2.5 Statistical analysis

The primary outcome was the Fm-100 Hue test score, and the secondary outcomes were Ishihara’s colour blindness test score, CBC test score, CBC severity and Yu Ziping’s colour blindness test score. Shapiro-Wilk tests were used to check the distribution of data. Continuous variables were described using mean & standard deviation (normally distributed data) and median & inter-quartile range (non-parametric data). The treatment effect was analyzed using the Kruskal-Wallis test, followed by Dunn’s multiple comparison test. The effect of weeks of treatment was analyzed using the Friedman test followed by Wilcoxon matched-pairs signed-rank test. Bonferroni post-hoc adjustments were used for multiple comparisons. Analyses were performed using R 4.2.1 and P□<□0.05 was considered statistically significant.

#### 2.6 Quality control

The researchers responsible for collecting data were professionally trained. Epidata3.0 software was used for data entry by two researchers independently. Then, errors were identified by comparing the input results of the two researchers. Throughout the study, all patients were tested by the same researcher with the same test method.

## Results

### 1 Baseline characteristics

A total of 92 subjects were initially recruited. After excluding 12 patients, 80 were finally analyzed with a mean age of 28.5 years ([12.5) with male predominance (91%). The baseline characteristics of patients in different groups in the study are shown in Table 1.

**Table 1.** Baseline Characteristics.

| Category | Treatment 1 |  | Treatment 2 |  | Treatment 3 |  | Control |  | P |
| --- | --- | --- | --- | --- | --- | --- | --- | --- | --- |
|  | Mean | SD | Mean | SD | Mean | SD | Mean | SD |  |
| Age* | 18.0 | 23.0 | 23.0 | 16.0 | 25.0 | 15.3 | 39.5 | 10.5 | <0.001 |
| Male (n, %) | 17.0 | 100.0 | 20.0 | 95.2 | 20.0 | 90.9 | 15.0 | 75.0 | 0.056 |
| FM-100 | 1864.0 | 630.3 | 2487.0 | 471.6 | 2452.7 | 809.1 | 2824.3 | 832.4 | 0.001 |
| Ishihara | 82.2 | 21.8 | 66.8 | 19.3 | 58.2 | 27.2 | 44.8 | 34.6 | <0.001 |
| CBC score* | 160.0 | 156.0 | 276.0 | 116.0 | 228.0 | 182.0 | 150.0 | 168.5 | 0.033 |
| CBC severity* | 1.0 | 1.0 | 3.0 | 2.0 | 2.3 | 1.5 | 2.8 | 3.3 | 0.030 |
| Yu* | 1.0 | 2.0 | 1.0 | 3.0 | 1.0 | 1.0 | 1.0 | 3.3 | 0.270 |
| *non-parametric |  |  |  |  |  |  |  |  |  |

### 2 Effect of treatments (each test score or each time point)

All 80 subjects underwent a complete 4-week treatment course and the corresponding examinations. The examination results at baseline and at the end of week 1, week 2 and week 4 of treatment are shown in the supplementary appendix 3.

After four weeks of treatment, the change in the FM-100 test score in treatment 1(Δ = 274 [283]), 2 (Δ = 316 [254]), and 3 (Δ = 340.5 [405.6])groups showed significant differences compared to the control group (Δ = 31 [226.25]; p =0.025, p =0.003, p < =0.002, respectively). In contrast,

There was no significant difference among the three treatment groups.

Ishihara’s test scores following treatment 2 (Δ = -9 [14]) were significantly lower than the control group (Δ = -1.5 [10.5], p =0.034) after two weeks of intervention. After four weeks of treatment, Ishihara’s test scores for treatment 2 (Δ = -17 [16]) and treatment 3 (Δ = -9 [16.3]) groups were significantly lower than the control group (Δ = 0.5 [10], p <0.001, p =0.21, respectively). Moreover, there was a significant difference between treatments 2 and 1 (Δ = -3 [11], p =0.048).

There was a significant difference in the change in CBC score among the treatment 2, 3 and control groups after two and four weeks of intervention (p < 0.001), with significantly lower scores observed in treatment 2 (Δ = -80 [92], Δ = -104 [118], respectively) and treatment 3 groups (Δ = -46 [100] and Δ = -64 [112.5], respectively) compared to the control group (Δ = 11 [45.5] and Δ = 4 [39], respectively; p <0.001, p =0.001, respectively). However, there was no difference among the four groups after one week (p = 0.071).

After two weeks of intervention, the change in CBC severity score of treatment 2 (Δ = 3 [3]) was significantly larger than the control group (Δ = 0 [2]; p = 0.006). After four weeks of treatment, the change in CBC severity score for treatment 2 (Δ = 3 [3]) and treatment 3 (Δ = 3 [3]) groups were significantly higher than the control group (Δ = 0 [1]; p < 0.001, p < 0.001, respectively) and treatment 1 group (Δ = 2 [3]; p = 0.029, p =0.017, respectively). As for Yu’s test score, the treatment 2 group (Δ = 3 [2], Δ = 5 [5] and Δ = 7 [5], respectively) showed a larger increase than the control group (Δ = 0 [1], Δ = 0 [1.3] and Δ = 0 [1], respectively) as from week 1 (p < 0.001, p < 0.001 and p < 0.001, respecitvely). The change in Yu’s test scores in treatment 1 (Δ = 2 [6] and Δ = 4 [6], respectively), and 3 groups (Δ = 3 [2.8] and Δ = 4 [5.5], respectively) were all larger than the control group after two and four weeks of intervention (treatment 1: p = 0.002, p = 0.004; treatment 3: p < 0.001, p < 0.001). There was no significant difference in Yu’s test scores among the three treatment groups.

### 3 Within treatment (comparison across time)

For the treatment 1 group, the FM-100 test score, CBC severity score and Yu’s test score showed significant increases from two and four weeks of treatment compared to baseline (FM-100 test score: p=0.014, p < 0.001; CBC severity score: p =0.028, p =0.04; Yu’s test score: p =0.03, p =0.02). The CBC scores significantly decreased from two and four weeks of treatment (p =0.016, p =0.006, respectively). There was a significant change in the FM-100 test score for treatment 1 as from week 1 (p =0.006). Ishihara’s test scores were similar for the treatment 1 group during the trial (p = 0.433). The FM-100 test score and Ishihara’s test scores for the treatment 2 group significantly differed from their pre-treatment score from two and four weeks of the treatment (FM-100 test score: p =0.01, p =0.002; Ishihara’s test score: p =0.035, p =0.006), while the other three test scores showed significant differences compared to baseline at 1, 2 and 4 weeks of intervention (CBC score: p =0.05, p < 0.001, p < 0.001; CBC severity score: p =0.007, p < 0.001, p < 0.001; Yu’s test score: p < 0.001, p < 0.001, p < 0.001).

As for the treatment 3 group, only Ishihara’s test scores showed a significant difference after four weeks of intervention compared to baseline (p =0.012), while there were significant changes in the other test scores at 1, 2 and 4 weeks of treatment (FM-100 test score: p < =0.008, p < 0.001, p < 0.001; CBC score: p =0.016, p =0.022, p =0.006; CBC severity score: p =0.003, p =0.004, p < 0.001; Yu’s test score: p=0.002, p < 0.001, p < 0.001).

## Discussion

Colour vision is an important visual function for people to recognize colours and is based on the differences in the sensitivity of L-cones, M-cones and S-cones in the retina to long-wave (557 nm; red), medium-wave (530 nm; green) and short-wave (426 nm; blue) light, respectively^[9, 10]^. CVD is caused by the decreased or poor ability of the cones on the retina to receive and distinguish different wavelengths of visible light^[11]^. According to the cause of the disease, congenital CVD can be divided into three categories: anomalous trichromat, dichromatic vision and achromatopsia. Anomalous trichromat is the most common colour vision deficiency^[12-14]^, which mainly includes congenital deficiency and acquired deficiency and is usually caused by L-cone or M-cone abnormality. In some cases, patients cannot distinguish between red and green, termed red-green colour vision defect^[1]^. There are more than 300 million CVD patients worldwide, and the prevalence rate of CVD in Asia is about 5% (8% in males and 0.4-0.8% in females) ^[2, 15]^. Patients that cannot distinguish between specific colours exhibit a limited ability to recognize objects in daily life^[16, 17]^. Steward et al. reported that the career choice of 34% of CVD patients was affected by their condition, and 24% were banned from specific occupations such as police, military, railway, electronics and communications^[18, 19]^. In addition, CVD patients may be subject to discrimination and further develop psychological disorders^[10]^. A case-control study found that stereoscopic vision decreased in CVD patients compared with healthy controls^[20]^. A related study indicated that the prevalence of migraine in male adolescents with CVD was 23% higher than in the control group^[21]^. Therefore, exploring effective intervention and treatment measures for CVD patients is of great clinical significance.

CVD is a congenital X-linked recessive disorder. To our knowledge, at present, there is no effective treatment for CVD. We used PBM to perform exploratory non-invasive physiotherapy for CVD patients and observed comparable therapeutic effects. Most importantly, this is the first study to substantiate that CVD patients can be effectively treated by non-invasive methods.

The results of our study suggested that the colour perception of CVD patients may be improved to some extent after receiving PBM treatment. The main outcome (the FM-100 test score) significantly improved from the second week, and the improvement was most significant at the end of the fourth week. At the same time, secondary outcomes, including Ishihara’s scores, CBC score, CBC severity and Yu Ziping’s test scores, were significantly improved from week 2. Ishihara’s colour blind book is an international colour blindness test book. Although Yu Ziping’s colour blind book is not internationally used, it is the most widely used colour blindness test book in China. Herein, we simultaneously applied these two kinds of colour blindness to detect the abnormal colour vision of patients and observed consistent evaluation results.

Currently, there are two main therapies for CVD; colour vision abnormalities of CVD patients can be corrected by using lenses made of special materials^[1, 22-25]^, while gene therapy involves the intravitreal injection of adenoviruses containing normal visual genes into the retina for individuals with CNGB3, GNAT2, M-opsin and L-opsin mutations. However, it should be borne in mind that optical lenses cannot fundamentally correct abnormal colour vision of CVD patients, and there are also certain risks associated with gene therapy. Compared with the above two treatments, PBM has gained significant attention, given its non-invasiveness and can effectively improve the abnormal colour vision of CVD patients, showing significant effectiveness and safety.

Animal studies have shown that exposure to light at 650-900nm wavelengths for several weeks may increase intracellular ATP and further improve mitochondrial function^[26, 27]^. It is speculated that PBM may affect the colour vision function of CVD patients by improving the mitochondrial function of cones. A recent study found that colour vision tests under different illuminances could affect the performance of CVD patients^[28]^. It has been suggested that illuminance could affect the colour discrimination of CVD patients.

The intervention method adopted in this study for patients with CVD is unprecedented, innovative and scientific. However, the sample size included in this study was relatively small, and the subjects were not followed up regularly after stopping the treatment. Accordingly, we failed to objectively evaluate whether PBM can cure CVD or can only provide short-term remission to patients. Our findings warrant further validation in longitudinal studies with larger sample sizes and longer follow-up duration.

In conclusion, PBM in patients with CVD yields a definite therapeutic effect, broadening this patient population’s therapeutic landscape.

## Data Availability

All data produced in the present study are available upon reasonable request to the authors.

## Data sharing statement

All relevant data are available within the manuscript.

## Contributors

Nandi Bao: Data Curation, Data Analysis, Writing - Original draft. Liang Jia: Study Design, Data Collection, Writing - Review and editing.

