## supplementary appendix 2 for "Novel non-invasive physical photobiomodulation can treat congenital colour vision deficiency and enhance color vision recognition ability: a randomized, single blind, controlled clinical trial"

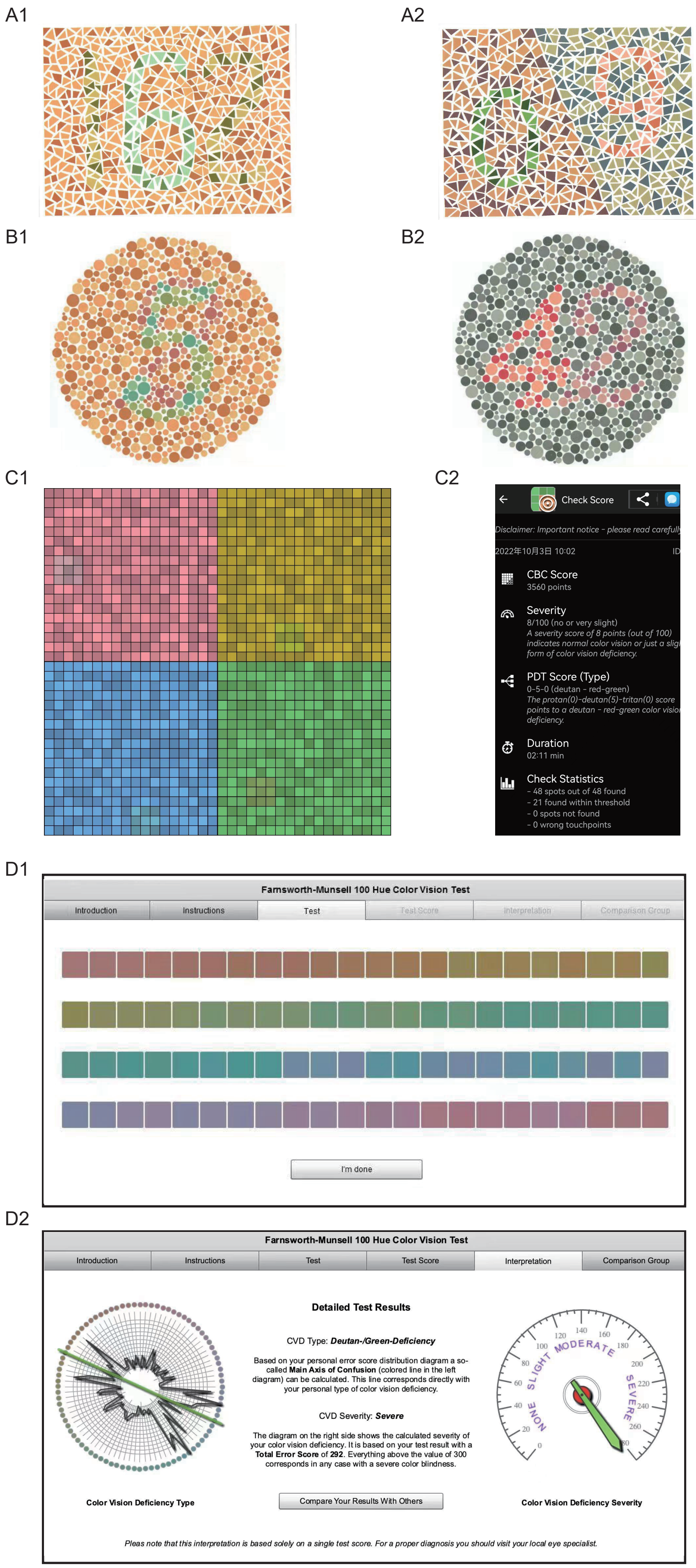


Supplementary appendix 2: Figure A1 and A2 are part of the color blindness pictures in Yu Ziping Color Blindness (People's Medical Publishing House, 6th Edition); Figure B1 and B2 are partial pictures of Ishihara Color blindness in Japan. Figure C1 shows the four basic screens of Color Blindness Check software for color vision detection. The color blocks with the same lightness and specific colors appear randomly in the uniform background noise, and deepen with time saturation until recognized and clicked by the patient; Figure C2 shows a screenshot of Color Blindness Check results: color vision score, severity, type of defect (red or green defect), and detection time. Figure D1 shows the color block arrangement matrix of FM-100 color phase detection. Figure D2 shows the results of FM-100 hue detection: hue confusion axis (red or green), total error score and severity.
