## supplementary appendix 3 for "Novel non-invasive physical photobiomodulation can treat congenital colour vision deficiency and enhance color vision recognition ability: a randomized, single blind, controlled clinical trial"

**Table 1: The examination results before and at the end of week 1, week 2 and week 4 of treatment group 1.**

| **Number** | **Gender** | **Type of CVD** | **FM-100** | | | | **Ishihara's test score** | | | | **CBC test score** | | | | **CBC severity** | | | | **YU’s test score** | | | |
| --- | --- | --- | --- | --- | --- | --- | --- | --- | --- | --- | --- | --- | --- | --- | --- | --- | --- | --- | --- | --- | --- | --- |
|  |  |  | **W0** | **W1** | **W2** | **W4** | **W0** | **W1** | **W2** | **W4** | **W0** | **W1** | **W2** | **W4** | **W0** | **W1** | **W2** | **W4** | **W0** | **W1** | **W2** | **W4** |
| 1 | male | Red | 52 | 32 | 56 | 60 | 3 | 5 | 1 | 2 | 2091 | 2452 | 2414 | 2307 | 80 | 64 | 71 | 66 | 0 | 12 | 7 | 11 |
| 2 | male | Red | 120 | 112 | 105 | 102 | 0 | 1 | 2 | 3 | 1577 | 1919 | 1978 | 2075 | 96 | 84 | 82 | 81 | 0 | 4 | 5 | 6 |
| 3 | male | Red | 416 | 405 | 379 | 367 | 0 | 0 | 0 | 0 | 1505 | 1111 | 1010 | 1352 | 95 | 100 | 98 | 99 | 0 | 0 | 0 | 1 |
| 4 | male | Red | 204 | 264 | 236 | 168 | 1 | 2 | 2 | 2 | 1254 | 1698 | 1626 | 1799 | 94 | 92 | 96 | 94 | 0 | 0 | 1 | 0 |
| 5 | male | Red | 152 | 135 | 104 | 96 | 0 | 1 | 1 | 2 | 1642 | 1794 | 2012 | 2085 | 97 | 92 | 91 | 87 | 0 | 0 | 0 | 1 |
| 6 | male | Red | 260 | 252 | 212 | 288 | 1 | 1 | 2 | 1 | 1639 | 1807 | 1816 | 1854 | 94 | 80 | 91 | 93 | 1 | 4 | 2 | 6 |
| 7 | male | Red | 160 | 136 | 136 | 136 | 1 | 2 | 3 | 4 | 1195 | 1743 | 1995 | 1995 | 98 | 91 | 91 | 91 | 1 | 7 | 8 | 8 |
| 8 | male | Red | 356 | 348 | 320 | 276 | 1 | 1 | 0 | 1 | 1755 | 1780 | 1865 | 1984 | 80 | 78 | 76 | 70 | 1 | 1 | 2 | 1 |
| 9 | male | Red | 220 | 120 | 150 | 120 | 1 | 2 | 2 | 3 | 1521 | 1986 | 1998 | 2094 | 94 | 100 | 99 | 95 | 0 | 1 | 2 | 3 |
| 10 | male | Red | 544 | 200 | 260 | 140 | 1 | 4 | 8 | 3 | 3715 | 3707 | 3803 | 3705 | 9 | 8 | 3 | 6 | 2 | 2 | 4 | 5 |
| 11 | male | Red | 592 | 510 | 400 | 376 | 2 | 2 | 2 | 0 | 1381 | 1409 | 1457 | 1641 | 93 | 93 | 95 | 96 | 0 | 0 | 0 | 1 |
| 12 | male | Red | 104 | 132 | 124 | 92 | 2 | 2 | 4 | 5 | 2681 | 2797 | 3018 | 2955 | 57 | 56 | 47 | 54 | 2 | 6 | 10 | 11 |
| 13 | male | Red | 52 | 48 | 28 | 52 | 1 | 1 | 4 | 4 | 1685 | 1793 | 1964 | 2070 | 89 | 89 | 97 | 93 | 8 | 9 | 12 | 12 |
| 14 | male | Red | 60 | 58 | 62 | 48 | 2 | 4 | 5 | 5 | 2010 | 2278 | 2158 | 2103 | 76 | 62 | 84 | 77 | 4 | 16 | 18 | 18 |
| 15 | male | Red | 60 | 64 | 56 | 50 | 4 | 0 | 6 | 6 | 2163 | 2320 | 2067 | 2227 | 78 | 80 | 81 | 62 | 5 | 13 | 15 | 17 |
| 16 | male | Red | 188 | 120 | 96 | 132 | 1 | 0 | 4 | 0 | 1406 | 1590 | 1570 | 1959 | 91 | 94 | 94 | 92 | 2 | 1 | 4 | 3 |
| 17 | male | Red | 152 | 138 | 120 | 116 | 1 | 0 | 1 | 1 | 2468 | 2539 | 2715 | 2755 | 76 | 67 | 56 | 58 | 0 | 2 | 3 | 5 |

**Table 2: The examination results before and at the end of week 1, week 2 and week 4 of treatment group 2.**

| **Number** | **Gender** | **Type of CVD** | **FM-100** | | | | | | | **Ishihara's test score** | | | | | | | **CBC test score** | | | | | | | **CBC severity** | | | | | | **YU’s test score** | | | | | | |
| --- | --- | --- | --- | --- | --- | --- | --- | --- | --- | --- | --- | --- | --- | --- | --- | --- | --- | --- | --- | --- | --- | --- | --- | --- | --- | --- | --- | --- | --- | --- | --- | --- | --- | --- | --- | --- |
|  |  |  | **W0** | **W1** | | **W2** | | **W4** | | **W0** | | **W1** | | **W2** | | **W4** | **W0** | **W1** | | **W2** | | **W4** | | **W0** | **W1** | | **W2** | | **W4** | **W0** | | **W1** | | **W2** | | **W4** |
| 1 | male | Green | 316 | | 278 | | 258 | | 218 | 13 | 15 | | 16 | | 17 | | 1912 | | 2064 | | 2167 | | 2296 | 79 | 79 | 73 | | 70 | | 1 | 9 | | 11 | | 13 | |
| 2 | male | Green | 176 | | 96 | | 86 | | 72 | 4 | 5 | | 6 | | 9 | | 2552 | | 2879 | | 3096 | | 3298 | 63 | 54 | 38 | | 28 | | 3 | 7 | | 5 | | 11 | |
| 3 | male | Green | 280 | | 104 | | 100 | | 90 | 3 | 4 | | 6 | | 7 | | 1863 | | 2406 | | 2531 | | 2671 | 71 | 81 | 75 | | 70 | | 0 | 5 | | 8 | | 9 | |
| 4 | male | Green | 160 | | 154 | | 142 | | 120 | 2 | 3 | | 3 | | 3 | | 2076 | | 2627 | | 2689 | | 2741 | 87 | 66 | 64 | | 60 | | 0 | 2 | | 3 | | 4 | |
| 5 | male | Green | 276 | | 316 | | 148 | | 176 | 2 | 5 | | 7 | | 6 | | 2092 | | 2499 | | 2854 | | 2968 | 78 | 64 | 58 | | 53 | | 0 | 10 | | 13 | | 17 | |
| 6 | female | Green | 316 | | 252 | | 218 | | 174 | 6 | 6 | | 6 | | 8 | | 2921 | | 3169 | | 3251 | | 3391 | 48 | 37 | 35 | | 30 | | 6 | 10 | | 13 | | 15 | |
| 7 | male | Green | 364 | | 192 | | 240 | | 120 | 3 | 2 | | 3 | | 3 | | 2743 | | 2424 | | 2305 | | 2767 | 60 | 74 | 76 | | 51 | | 1 | 4 | | 3 | | 2 | |
| 8 | male | Green | 296 | | 184 | | 124 | | 138 | 3 | 5 | | 20 | | 18 | | 2375 | | 2293 | | 3437 | | 3379 | 74 | 68 | 25 | | 31 | | 1 | 3 | | 15 | | 13 | |
| 9 | male | Green | 304 | | 180 | | 120 | | 124 | 3 | 6 | | 5 | | 5 | | 2083 | | 1754 | | 2346 | | 2399 | 80 | 79 | 71 | | 63 | | 1 | 1 | | 1 | | 2 | |
| 10 | male | Green | 236 | | 220 | | 187 | | 174 | 3 | 3 | | 7 | | 5 | | 2640 | | 2698 | | 3173 | | 2967 | 66 | 57 | 36 | | 43 | | 0 | 4 | | 7 | | 6 | |
| 11 | male | Green | 112 | | 124 | | 140 | | 106 | 2 | 3 | | 4 | | 5 | | 2159 | | 2203 | | 2315 | | 2461 | 86 | 84 | 81 | | 75 | | 0 | 3 | | 3 | | 6 | |
| 12 | male | Green | 356 | | 288 | | 228 | | 160 | 1 | 1 | | 4 | | 4 | | 2465 | | 2555 | | 2626 | | 2727 | 71 | 68 | 63 | | 52 | | 1 | 1 | | 2 | | 1 | |
| 13 | male | Green | 132 | | 88 | | 96 | | 124 | 0 | 6 | | 5 | | 15 | | 3263 | | 3597 | | 3607 | | 3479 | 24 | 8 | 11 | | 13 | | 5 | 11 | | 11 | | 19 | |
| 14 | male | Green | 308 | | 408 | | 276 | | 212 | 1 | 4 | | 6 | | 6 | | 1906 | | 2039 | | 2101 | | 2208 | 87 | 76 | 74 | | 70 | | 0 | 2 | | 3 | | 4 | |
| 15 | male | Green | 228 | | 180 | | 148 | | 104 | 6 | 7 | | 7 | | 6 | | 3389 | | 3447 | | 3664 | | 3725 | 20 | 17 | 13 | | 4 | | 1 | 3 | | 6 | | 8 | |
| 16 | male | Green | 344 | | 440 | | 384 | | 356 | 3 | 5 | | 6 | | 6 | | 3054 | | 3157 | | 3489 | | 3489 | 58 | 47 | 21 | | 21 | | 3 | 6 | | 7 | | 10 | |
| 17 | male | Green | 204 | | 156 | | 100 | | 76 | 6 | 6 | | 7 | | 7 | | 2787 | | 2647 | | 2642 | | 2932 | 70 | 69 | 58 | | 45 | | 4 | 7 | | 12 | | 13 | |
| 18 | male | Green | 312 | | 180 | | 156 | | 148 | 3 | 3 | | 6 | | 12 | | 2269 | | 2102 | | 2202 | | 2230 | 72 | 83 | 85 | | 86 | | 2 | 7 | | 11 | | 16 | |
| 19 | male | Green | 196 | | 186 | | 160 | | 200 | 1 | 2 | | 5 | | 3 | | 2084 | | 2128 | | 2190 | | 2346 | 84 | 84 | 75 | | 76 | | 4 | 5 | | 5 | | 8 | |
| 20 | male | Green | 136 | | 156 | | 120 | | 120 | 2 | 2 | | 6 | | 8 | | 2430 | | 2557 | | 2509 | | 2547 | 82 | 71 | 76 | | 72 | | 1 | 4 | | 6 | | 8 | |
| 21 | male | Green | 216 | | 160 | | 136 | | 112 | 5 | 5 | | 6 | | 8 | | 3165 | | 2706 | | 2655 | | 2562 | 42 | 60 | 63 | | 69 | | 2 | 6 | | 6 | | 8 | |

**Table 3:** **The examination results before and at the end of week 1, week 2 and week 4 of treatment group 3.**

| **Number** | **Gender** | **Type of CVD** | **FM-100** | | | | | | | **Ishihara's test score** | | | | | | **CBC test score** | | | | | | | **CBC severity** | | | | | **YU’s test score** | | | | | | |
| --- | --- | --- | --- | --- | --- | --- | --- | --- | --- | --- | --- | --- | --- | --- | --- | --- | --- | --- | --- | --- | --- | --- | --- | --- | --- | --- | --- | --- | --- | --- | --- | --- | --- | --- |
|  |  |  | **W0** | **W1** | | **W2** | | **W4** | | **W0** | | **W1** | | **W2** | **W4** | **W0** | **W1** | | **W2** | | **W4** | | **W0** | **W1** | | **W2** | **W4** | **W0** | | **W1** | | **W2** | | **W4** |
| 1 | male | Red-Green | 376 | | 232 | | 200 | | 244 | 3 | 7 | | 9 | | 12 | 2076 | | 2593 | | 2724 | | 2671 | 71 | 62 | 62 | | 61 | 2 | 6 | | 8 | | 13 | |
| 2 | male | Red-Green | 712 | | 532 | | 624 | | 516 | 0 | 1 | | 0 | | 3 | 675 | | 800 | | 714 | | 810 | 100 | 100 | 100 | | 100 | 0 | 4 | | 3 | | 5 | |
| 3 | male | Red-Green | 512 | | 328 | | 300 | | 232 | 0 | 4 | | 3 | | 5 | 2328 | | 2176 | | 2150 | | 2405 | 65 | 75 | 73 | | 73 | 3 | 1 | | 3 | | 6 | |
| 4 | male | Red-Green | 148 | | 140 | | 132 | | 112 | 3 | 5 | | 7 | | 11 | 3482 | | 3614 | | 3748 | | 3623 | 18 | 11 | 6 | | 8 | 2 | 3 | | 8 | | 11 | |
| 5 | female | Red-Green | 192 | | 120 | | 72 | | 104 | 3 | 7 | | 2 | | 4 | 3271 | | 3380 | | 3283 | | 3516 | 28 | 20 | 27 | | 20 | 0 | 2 | | 2 | | 8 | |
| 6 | male | Red-Green | 332 | | 212 | | 264 | | 176 | 1 | 4 | | 1 | | 6 | 1936 | | 2222 | | 2304 | | 2318 | 78 | 81 | 76 | | 76 | 1 | 3 | | 2 | | 3 | |
| 7 | male | Red-Green | 192 | | 200 | | 180 | | 138 | 1 | 3 | | 4 | | 5 | 3333 | | 3126 | | 3298 | | 3496 | 28 | 39 | 30 | | 25 | 0 | 0 | | 1 | | 2 | |
| 8 | male | Red-Green | 408 | | 236 | | 240 | | 292 | 1 | 1 | | 3 | | 3 | 1547 | | 1971 | | 1965 | | 2018 | 87 | 83 | 79 | | 88 | 0 | 0 | | 4 | | 2 | |
| 9 | male | Red-Green | 1096 | | 508 | | 372 | | 436 | 3 | 4 | | 6 | | 6 | 1668 | | 1975 | | 2467 | | 2475 | 89 | 88 | 74 | | 78 | 1 | 1 | | 4 | | 3 | |
| 10 | male | Red-Green | 292 | | 560 | | 488 | | 436 | 0 | 5 | | 6 | | 7 | 1944 | | 2096 | | 2178 | | 2282 | 75 | 80 | 81 | | 83 | 0 | 4 | | 3 | | 2 | |
| 11 | male | Red-Green | 316 | | 298 | | 272 | | 256 | 4 | 4 | | 3 | | 6 | 1642 | | 1754 | | 2263 | | 2403 | 95 | 84 | 82 | | 68 | 1 | 4 | | 7 | | 7 | |
| 12 | male | Red-Green | 200 | | 156 | | 168 | | 196 | 5 | 5 | | 9 | | 7 | 3339 | | 3452 | | 3600 | | 3572 | 25 | 21 | 17 | | 20 | 3 | 7 | | 11 | | 15 | |
| 13 | male | Red-Green | 508 | | 496 | | 596 | | 436 | 5 | 7 | | 5 | | 7 | 3014 | | 3095 | | 3172 | | 3486 | 35 | 32 | 37 | | 12 | 1 | 3 | | 4 | | 6 | |
| 14 | male | Red-Green | 160 | | 132 | | 126 | | 118 | 5 | 3 | | 3 | | 6 | 3024 | | 3286 | | 3231 | | 3367 | 38 | 26 | 28 | | 21 | 1 | 1 | | 2 | | 3 | |
| 15 | male | Red-Green | 212 | | 184 | | 188 | | 192 | 6 | 6 | | 6 | | 6 | 2679 | | 2703 | | 2632 | | 2435 | 54 | 58 | 62 | | 66 | 3 | 4 | | 4 | | 4 | |
| 16 | male | Red-Green | 176 | | 116 | | 128 | | 128 | 1 | 2 | | 2 | | 1 | 2556 | | 2642 | | 2816 | | 2696 | 64 | 61 | 60 | | 61 | 3 | 4 | | 4 | | 5 | |
| 17 | male | Red-Green | 368 | | 132 | | 256 | | 160 | 1 | 3 | | 5 | | 6 | 1373 | | 1373 | | 1699 | | 1946 | 93 | 89 | 76 | | 66 | 1 | 2 | | 3 | | 4 | |
| 18 | male | Red-Green | 152 | | 140 | | 126 | | 108 | 1 | 2 | | 3 | | 4 | 3049 | | 3408 | | 3502 | | 3584 | 51 | 29 | 20 | | 21 | 3 | 4 | | 6 | | 13 | |
| 19 | female | Red-Green | 244 | | 160 | | 56 | | 88 | 4 | 4 | | 5 | | 7 | 2327 | | 2886 | | 2875 | | 2885 | 63 | 55 | 50 | | 44 | 1 | 5 | | 4 | | 10 | |
| 20 | male | Red-Green | 92 | | 148 | | 152 | | 126 | 2 | 5 | | 6 | | 7 | 3510 | | 3510 | | 3709 | | 3798 | 24 | 25 | 10 | | 8 | 1 | 4 | | 5 | | 6 | |
| 21 | male | Red-Green | 208 | | 244 | | 204 | | 284 | 1 | 4 | | 5 | | 9 | 1844 | | 2219 | | 2304 | | 2565 | 79 | 91 | 82 | | 66 | 1 | 4 | | 6 | | 7 | |
| 22 | male | Red-Green | 204 | | 120 | | 128 | | 136 | 1 | 3 | | 5 | | 7 | 3342 | | 3273 | | 3310 | | 3333 | 21 | 31 | 29 | | 23 | 1 | 4 | | 4 | | 3 | |

**Table 4: The examination results before and at the end of week 1, week 2 and week 4 of control group.**

| **Number** | **Gender** | **Type of CVD** | **FM-100** | | | | **Ishihara's test score** | | | | **CBC test score** | | | | **CBC severity** | | | | | **YU’s test score** | | | |
| --- | --- | --- | --- | --- | --- | --- | --- | --- | --- | --- | --- | --- | --- | --- | --- | --- | --- | --- | --- | --- | --- | --- | --- |
|  |  |  | **W0** | **W1** | **W2** | **W4** | **W0** | **W1** | **W2** | **W4** | **W0** | **W1** | **W2** | **W4** | | **W0** | **W1** | **W2** | **W4** | **W0** | **W1** | **W2** | **W4** |
| 1 | female | Red-Green | 80 | 82 | 96 | 85 | 14 | 13 | 14 | 15 | 3817 | 3802 | 3706 | 3825 | | 6 | 6 | 5 | 6 | 12 | 13 | 12 | 13 |
| 2 | male | Red | 312 | 334 | 350 | 325 | 0 | 0 | 0 | 0 | 2080 | 2005 | 1958 | 1955 | | 81 | 83 | 86 | 91 | 0 | 0 | 0 | 0 |
| 3 | male | Red | 148 | 256 | 208 | 232 | 3 | 4 | 5 | 4 | 3482 | 3555 | 3639 | 3614 | | 18 | 17 | 9 | 11 | 2 | 5 | 2 | 3 |
| 4 | female | Red-Green | 60 | 12 | 28 | 38 | 10 | 10 | 6 | 8 | 3682 | 3313 | 3756 | 3459 | | 4 | 20 | 4 | 14 | 10 | 18 | 17 | 17 |
| 5 | male | Red-Green | 68 | 80 | 96 | 88 | 7 | 7 | 7 | 6 | 3594 | 3482 | 3574 | 3501 | | 10 | 17 | 14 | 15 | 5 | 6 | 5 | 6 |
| 6 | male | Red | 86 | 88 | 84 | 80 | 5 | 7 | 5 | 4 | 3816 | 3796 | 3838 | 3805 | | 2 | 13 | 6 | 11 | 1 | 1 | 1 | 2 |
| 7 | male | Red | 128 | 134 | 130 | 124 | 1 | 1 | 0 | 1 | 1877 | 2095 | 1978 | 2086 | | 91 | 80 | 85 | 86 | 1 | 1 | 0 | 1 |
| 8 | male | Red | 148 | 120 | 134 | 128 | 0 | 0 | 0 | 0 | 2154 | 2374 | 2259 | 2309 | | 82 | 78 | 80 | 79 | 0 | 1 | 1 | 1 |
| 9 | male | Red | 320 | 276 | 356 | 338 | 1 | 1 | 1 | 1 | 1399 | 1663 | 1564 | 1621 | | 99 | 92 | 91 | 95 | 1 | 1 | 0 | 1 |
| 10 | male | Green | 188 | 136 | 200 | 192 | 9 | 7 | 8 | 8 | 3851 | 3804 | 3892 | 3824 | | 3 | 6 | 0 | 4 | 12 | 12 | 10 | 11 |
| 11 | female | Red-Green | 244 | 148 | 180 | 208 | 4 | 9 | 5 | 4 | 2327 | 2510 | 2772 | 2676 | | 63 | 61 | 51 | 58 | 6 | 6 | 5 | 5 |
| 12 | male | Green | 92 | 144 | 104 | 126 | 2 | 2 | 2 | 2 | 3510 | 3523 | 3569 | 3512 | | 24 | 17 | 19 | 18 | 1 | 1 | 1 | 1 |
| 13 | female | Red-Green | 80 | 96 | 82 | 104 | 0 | 0 | 1 | 0 | 3526 | 3480 | 3727 | 3675 | | 16 | 20 | 7 | 9 | 0 | 0 | 1 | 1 |
| 14 | male | Green | 208 | 196 | 218 | 224 | 1 | 0 | 5 | 4 | 1844 | 2033 | 2265 | 2198 | | 79 | 88 | 85 | 87 | 1 | 3 | 3 | 3 |
| 15 | male | Red-Green | 304 | 304 | 356 | 348 | 4 | 5 | 6 | 5 | 2314 | 1545 | 1848 | 1678 | | 59 | 87 | 77 | 83 | 4 | 4 | 3 | 4 |
| 16 | male | Green | 136 | 196 | 166 | 140 | 2 | 6 | 4 | 4 | 2266 | 1753 | 1894 | 1950 | | 69 | 84 | 75 | 73 | 4 | 3 | 4 | 3 |
| 17 | male | Green | 760 | 630 | 830 | 736 | 1 | 1 | 3 | 2 | 2647 | 2718 | 2519 | 2601 | | 51 | 53 | 67 | 56 | 1 | 1 | 0 | 1 |
| 18 | female | Green | 152 | 150 | 138 | 148 | 3 | 4 | 2 | 3 | 3243 | 3214 | 3362 | 3297 | | 29 | 35 | 23 | 31 | 3 | 3 | 3 | 3 |
| 19 | male | Green | 452 | 412 | 274 | 374 | 0 | 2 | 2 | 1 | 1714 | 1801 | 2240 | 2103 | | 89 | 94 | 82 | 87 | 1 | 1 | 0 | 1 |
| 20 | male | Red-Green | 204 | 164 | 136 | 140 | 1 | 3 | 5 | 4 | 3342 | 3504 | 3407 | 3468 | | 21 | 16 | 20 | 19 | 1 | 3 | 3 | 3 |
